# Proteomic biomarkers of cognitive function, *APOE* ε4 status, and dementia in Generation Scotland

**DOI:** 10.64898/2026.01.27.26344912

**Authors:** Hannah M. Smith, Joanna E. Moodie, Gail Davies, Anne Richmond, Josephine A. Robertson, Aleksandra D. Chybowska, Camilla Drake, Poppy Adkin, Spyros I. Vernardis, Arturas Grauslys, Matthew E. H. White, Sergej Andrejev, Charles Brigden, Christoph B. Messner, Caroline Hayward, Aleksej Zelezniak, Markus Ralser, Ilse Krätschmer, Matthew R. Robinson, Simon R. Cox, Riccardo E. Marioni

## Abstract

Untargeted mass spectrometry remains underutilised for blood-based biomarker discovery in dementia research from large cohorts, where affinity-based approaches dominate. To address this, we examined mass-spectrometry–derived proteomic correlates of cognitive function, genetic predisposition to cognitive health, *APOE* ε4 status, and incident dementia. Using multivariate Bayesian regression, we assessed associations between 439 independent mass spectrometry signals and five cognitive outcomes—four cognitive tests and a cognitive function polygenic score—in Generation Scotland (n = 14,258). We observed associations between three cognitive tests (digit symbol, vocabulary and verbal fluency) and a mass spectrometry signal that mapped to selenoprotein P (absolute β_range_ = 0.02 – 0.03, posterior inclusion probability ≥ 0.95). Carrying one or two copies of the *APOE* ε4 allele was associated with a lower mass spectrometry signal that mapped to afamin (β = -0.08 and -0.2, respectively, p < 3.1 x 10^-4^, n = 14,745). A higher mass spectrometry signal that maps to both complement c2 and complement factor B proteins was associated with lower hazard for incident dementia (hazard ratio = 0.75, p < 3.1 x 10^-4^, n_cases_ = 212 and n_controls_ = 6,765) diagnosed up to 17 years after blood sampling. We identified specific independent mass spectrometry signals which may be candidate biomarkers of cognitive function, *APOE* ε4 status, and could aid in the early detection of dementia; however, further replication studies in other populations are required.

## Introduction

Dementia is an umbrella term encompassing several subtypes of cognitive disorders, including Alzheimer’s disease (AD), vascular dementia, Lewy body dementia and frontotemporal dementia. A hallmark of dementia is a decline in cognitive function, predominantly memory loss, with deficits in other cognitive domains ^1^. The current global estimate of dementia prevalence stands at ∼ 55 million and is expected to rise to 152 million by 2050 ^2^. Given the prevalence and increasing number of cases, identifying biological markers (biomarkers) of dementia for early detection and intervention is a high priority.

Blood-based biomarkers are of great interest due to the accessible nature of bio-sampling, and may help to prioritise those at greatest of dementia when they first report having memory impairments. Proteins that correlate with cognitive functioning or predict future dementia risk may also reveal biological processes contributing to cognitive ageing. Importantly, integrating proteomic correlates of both observed cognitive function and genetic predisposition to cognitive health distinguishes markers of current cognitive state and those that may reflect lifelong, inherited pathways influencing cognition. Several blood-based proteins have shown promise as biomarkers of dementia, including growth differentiation factor 15 (GDF15), neurofilament light chain (NfL), glial fibrillary acidic protein (GFAP), and phosphorylated tau ^3–5^, which are indicators of neurodegeneration, glial activation, and tau pathology. However, in large-scale biomarker discovery studies, targeted protein panels that measure a predefined subset of the proteome are typically used ^3,6^. This may limit the potential for identifying biomarkers or biological pathways relevant to cognitive ageing and dementia ^7^.

Recent advances in ultra-high-throughput mass spectrometry ^8^ now allow for untargeted measurement of circulating proteins at scale. Using this approach, 439 serum protein/protein group levels have been measured in > 14,000 participants of the Generation Scotland cohort. In this study, we use data from 14,258 individuals to examine associations between these protein levels and four cognitive tests—digit symbol, verbal fluency, vocabulary, and logical memory—as well as a cognitive polygenic score (PGS). Given that these outcomes are correlated, we employ a Bayesian multivariate multiple linear regression model, which accounts for covariance among proteins and cognitive traits to improve statistical power and control for false positive results.

We also test whether any mass spectrometry signals associate with *APOE* ε4 status, a major genetic risk factor for late-onset AD ^9^, using linear mixed-effects models (n = 14,745). If *APOE* ε4 carriers exhibit distinct proteomic profiles, this may provide insight into molecular pathways that precede or contribute to late-onset AD. Finally, we test for associations between mass spectrometry signals and incident all-cause dementia in Generation Scotland participants diagnosed up to 17 years after blood sampling (n_cases_ = 212 and n_controls_ = 6,765) via mixed effects Cox proportional hazards regression.

## Methods

### Generation Scotland cohort

Generation Scotland (GS) is a family-based study of 23,960 individuals between the ages of 17 and 99 years old, residing in Scotland at the time of recruitment between 2006-2011 ^10^. Genetic, epigenetic, proteomic, clinical, lifestyle and sociodemographic information are available for the participants of GS. All components of GS received ethical approval from the NHS Tayside Committee on Medical Research Ethics (REC reference no. 05/S1401/89). GS has also been granted Research Tissue Bank status by the East of Scotland Research Ethics Service (REC reference no. 20-ES-0021), providing generic ethical approval for a wide range of uses within medical research.

### Cognitive testing

Four cognitive tests were administered in GS. The Wechsler Adult Intelligence Scale III digit symbol test ^11^ was used to assess processing speed, and the Wechsler Memory Scale III logical memory test ^12^ was used to assess immediate and delayed recall. The language-based cognitive abilities of participants were assessed via the verbal fluency test ^13^ and the Mill Hill vocabulary test ^14^.

### Genetic data

Samples were genotyped using Illumina HumanOmniExpressExome-8v1 chip and the Beadstudio-Gencall v3 genotype calling algorithm. Quality control excluded individuals with a call rate ≤ 98% and single nucleotide polymorphisms (SNPs) with a call rate of ≤ 98%, Hardy-Weinberg equilibrium (HWE) of ≤ 1×10^-6^ and minor allele frequency (MAF) of ≤ 1%. Phasing of the genotyped SNPs was carried out using SHAPEIT2 ^15^, and imputation was performed using the Haplotype Reference Consortium reference panel (HRC.r1-1) ^16^ on the Sanger Imputation Server with the PBWT software ^17^. Following imputation, further quality control excluded duplicate and monomorphic SNPs and SNPs with an imputation quality score of < 0.4.

### Polygenic score (PGS)

A cognitive PGS was derived using summary statistics from a subset (GS data removed) of a published meta-analytic GWAS of general cognitive function ^18^. The following CHARGE cohorts requested that their data be removed due to restrictions on their use: The Aging Gene-Environment Susceptibility - Reykjavik Study (AGES), The Atherosclerosis Risk in Communities Study (ARIC), The Cardiovascular Health Study (CHS), The Framingham Heart Study (FHS), and The Genetic Epidemiology Network of Arteriopathy (GENOA). The new sample size of meta-analytic GWAS after these cohorts and GS were removed was 263,184 individuals. The pgsc_calc workflow (version 2) ^19^ was used to calculate the PGS in GS using the summary statistics previously mentioned. A higher cognitive PGS indicates a higher cognitive function score.

### Proteomics

The 439 blood-based mass spectrometry signals in 15,818 individuals were measured using an ultra-high-throughput mass spectrometry method ^8,20^. A detailed protocol has been published previously ^20,21^. Briefly, the proteins in the samples were denatured and treated with trypsin before analysis. The samples were analysed using liquid chromatography-mass spectrometry (Agilent 1290 Infinity II system and TripleTOF 6600 mass spectrometer (SCIEX)) with a scanning SWATH method ^8^. The resulting data were analysed using DIA-NN (version 1.8.12) ^22^. Identification was carried out using a project-independent public spectral library ^23^. A precursor false discovery rate (FDR) was set to 1%. R (version 4.3.1) was used for post-processing. The linear batch-correction function from the limma package (version 3.54.2) was used to correct for different batches ^24^. Universal Protein Resource (UniProt) was used to annotate the dataset ^25^. Of the 439 signals identified, 133 were uniquely mapped to one protein, 107 mapped to several different proteins and 199 mapped to multiple protein isoforms. Within the 199 signals, 138 signals related to proteins that were identified more than once. This is due to different combinations of isoform IDs being identified. For example, the protein Apolipoprotein D was identified twice within the 199 signals, as one signal mapped to three isoforms of Apolipoprotein D [Uniprot identifiers: P05090, C9JF17, C9JX71], and was thus named Apolipoprotein D (G1). Whilst another signal mapped only to two isoforms of Apolipoprotein D [Uniprot identifiers: P05090, C9JF17], this was named Apolipoprotein D (G2). The “GX” naming system, where “X” refers to a number, is used to identify these repeated isoform groups. The remaining 61 signals that mapped to protein isoform groups that did not repeat have a “G” only after their protein name. The “PG X” naming system is used to identify the 107 signals that map to multiple different proteins. For example, one signal mapped to Immunoglobulin lambda variable 3-27, 3-25 and 3-16, and has been named protein group 1 (PG 1). All of the gene names the signal maps to are also included in parentheses, e.g., “PG 1 (IGLV3-16, IGLV3-25, IGLV3-27)”. Protein/protein groups underwent inverse-rank based normal transformation prior to all downstream analysis.

### Covariates

The Lancet Commission 2024 on dementia prevention recently released an updated list of risk factors for dementia ^26^. In our analysis, we adjusted the cognitive, *APOE* ε4 and dementia models for those that were measured in GS, which included: body mass index (BMI; kg/m^2^), self-reported alcohol intake (units in the past week), self-reported depression status (yes/no), education (an ordinal variable with 11 categories: 0: 0 years, 1: 1-4 years, 2: 5-9 years, 3: 10-11 years, 4: 12-13 years, 5: 14-15 years, 6: 16-17 years, 7: 18-19 years, 8: 20-21 years, 9: 22-23 years, 10: 24+ years), self-reported high blood pressure (yes/no), self-report diabetes and an epigenetic smoking score (EpiSmokEr). EpiSmokEr was calculated from DNA methylation data using the smoking score (SSc) method from the EpiSmokEr R package ^27^. The SSc method uses weights for 187 DNA methylation sites found to be associated with smoking ^28^. Methylation levels at these sites are multiplied by the weights and then summed to derive the EpiSmokEr score. We also included deprivation status as measured by the Scottish Index of Multiple Deprivation (SIMD) and family structure (modelled with a kinship matrix constructed using the kinship2 R package [version: 1.9.6.1] ^29^). The dementia models were additionally adjusted for *APOE* ε4 allele count (0/1/2).

### Cognitive analysis

A multivariate multiple linear regression Bayesian joint sparse model was conducted using MAJA ^30^ to estimate effect sizes for 439 proteins/protein groups in relation to four cognitive tests (digit symbol, logical memory, verbal fluency and vocabulary) and a cognitive PGS in a single regression model (with five outcomes and 439 predictors). By jointly and conditionally estimating effect sizes, MAJA takes into consideration the correlation structure within the proteomic data, whilst also considering their shared and unique relationships with the five outcomes. MAJA uses a multivariate spike-slab prior distribution to allow for features (proteins) with zero effect sizes and those with non-zero effect sizes. Parameters are estimated using a Gibbs sampler, a Markov Chain Monte Carlo method. We specified 5,000 iterations with 1,000 burn-in. ‘Significant’ associations are defined as proteins with a posterior inclusion probability (PIP) ≥ 0.95, and where the 95% credible interval for the estimate did not include zero. In the basic-adjusted models (n = 14,577), proteins/protein groups were regressed on age, sex and family structure using linear mixed-effects models (lmekin function from coxme package in R version: 2.2.22) ^31^. In fully-adjusted models (n = 14,258), the proteins/protein groups were additionally regressed on SIMD, BMI, alcohol intake, EpiSmokEr, depression status, education, and high blood pressure status. The scaled (mean zero and unit variance) residuals from both models were used as predictors, and all outcomes were also scaled for the analysis. Missing covariates were imputed (using the KNN function in the R package VIM ^32^) for individuals with at least 40% of the covariates measured to maximise sample size. The percentage of missing values for each covariate was as follows: 9% for alcohol intake, 6% for SIMD, 3% for EpiSmokEr, 5% for education, 2% for self-reported depression, self-reported diabetes, and self-reported high blood pressure, and 1% for BMI. There was no missing data for age and sex. Descriptive statistics for the participants based on non-imputed data for the cognitive models can be observed in **Table S1**.

### *APOE* ε4 analysis

Linear mixed effects models were performed using the lmekin function from the coxme R package ^31^. Proteins were modelled as the outcome, and *APOE* ε4 allele count – modelled as a factor (zero, one or two copies) – was included as a predictor. Basic models (n = 15,030) adjusted for age, sex and relatedness were performed, followed by fully adjusted models (n = 14,745) additionally adjusting for SIMD, BMI, alcohol intake, EpiSmokEr, depression status, education, and high blood pressure status. As with the cognitive models, missing covariates were imputed for individuals with at least 40% of covariates measured. The percentage of missing values for each covariate was as follows: 9% for alcohol intake, 6% for SIMD, 3% for EpiSmokEr, 5% for education, 1% for BMI, and 2% for self-reported depression, self-reported diabetes, and self-reported high blood pressure. There was no missing data for age and sex. A principal component analysis (PCA) of the proteomic data was performed using the scikit-learn package in Python (2.7.17) ^33^. The number of principal components it took to explain 80% of the variance in the proteomic data was used to calculate a Bonferroni adjusted p-value threshold (i.e., 0.05/161 = 3.1 x 10^-4^). Associations were therefore considered statistically significant if p < 3.1 x 10^-4^. Descriptive statistics for the participants included in the fully adjusted *APOE* ε4 based on non-imputed data can be observed in **Table S2**.

### Dementia information and analysis

Dementia diagnosis information was obtained through linkage to primary care records for ∼40% of individuals (due to consent restrictions with the data holders – individual general practitioners [GP] surgeries) and secondary healthcare records for everyone up to October 2023. If an individual was not diagnosed with dementia, then the censor was set at their age in October 2023 or age at death if they died prior to October 2023 without being diagnosed with dementia. A filter of ≥ 65 years was applied to age at diagnosis for cases, or age at censor for controls. Mixed effects Cox proportional hazard models ^31^ were used to test associations between proteins/protein groups and time-to-dementia. Initially, models were run adjusting for age, sex and relatedness (basic model). Further models were performed, additionally adjusting for SIMD, BMI, alcohol intake, EpiSmokEr, depression status, education, high blood pressure status, and *APOE* ε4 allele count (full model). n_cases_ = 216 and n_controls_ = 6,911 were included in the models with basic adjustments, n_cases_ = 212 and n_controls_ = 6,765 were included in the models with full adjustments. Similar to the cognitive models, in the fully adjusted models, missing covariate data were imputed for individuals with at least 4 of the 10 covariates measured to maximise sample size. The percentage of missing values for each covariate was as follows: 10% for alcohol intake, 6% for SIMD, 6% for EpiSmokEr, 5% for education, 1% for self-reported depression, self-reported diabetes, self-reported high blood pressure and BMI. There was no missing data for age and sex. A kinship matrix (described above) was included as a random effect to account for family structure in GS in both basic and full models. The same PCA-based Bonferroni adjusted threshold (p < 3.1 x 10^-4^) as in the *APOE* ε4 analyses above was used to determine if associations were statistically significant. Descriptive statistics for the participants included in the fully adjusted dementia analysis based on non-imputed data can be observed in **Table S3**.

## Results

### Proteomic associations with cognitive function

The MAJA models adjusting for age, sex and relatedness, identified 10, 6, 7 and 10 proteins associated (PIP ≥ 0.95) with digit symbol, logical memory, verbal fluency and vocabulary test scores, respectively (**Figure S1, Table S4**). Additionally, 8 proteins were associated (PIP ≥ 0.95) with the cognitive PGS. In the fully adjusted models (including BMI, alcohol, SIMD, EpiSmokEr, self-report depression, self-report diabetes, high blood pressure status, and education), selenoprotein P (G) was the only protein associated with cognitive test scores, which included vocabulary, digit symbol and verbal fluency (absolute β_range_: 0.02 – 0.03, **Figure 1, Table S5**). No other proteins/protein groups were associated with cognitive tests in the full model, and no associations (PIP ≥ 0.95) were observed for the logical memory cognitive test. Complement C4-B (G), PG 52, PG 54 and PG 56 were associated with the cognitive PGS (β = 0.08, -0.09, 0.08 and -0.08, respectively).

**Figure 1:**
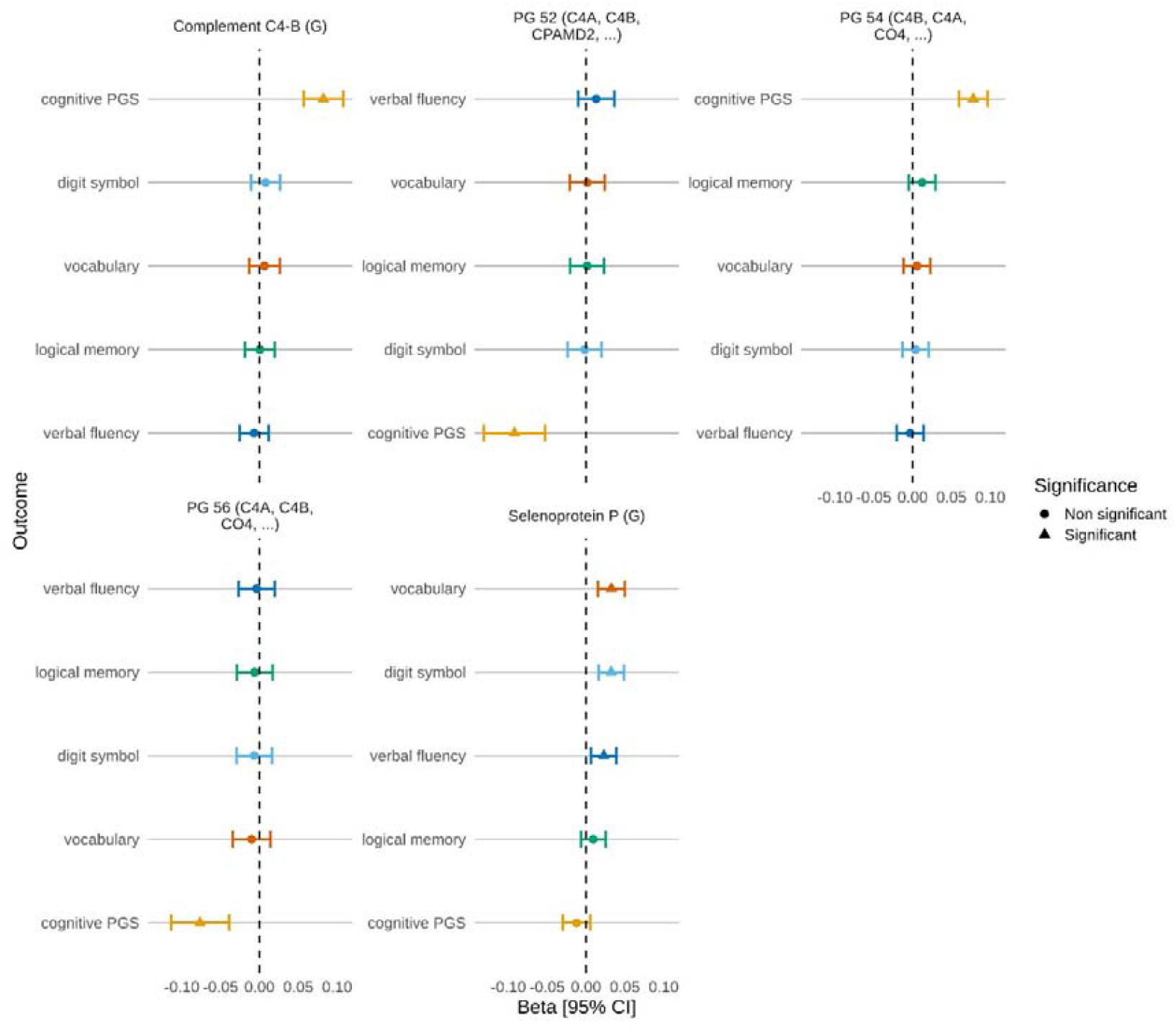
Associations between protein/protein group levels and four cognitive tests, and cognitive PGS in Generation Scotland. Standardised effect sizes (Beta) are shown for seven proteins/protein groups with digit symbol (light blue), logical memory (green), vocabulary (orange), verbal fluency (dark blue), and cognitive PGS (yellow) in Generation Scotland. Protein/protein groups are included in the plot if they had a significant association (PIP ≥ 0.95 and 95% credible interval did not cross zero) with one or more outcomes. Error bars represent the 95% credible interval [95% CI].

The estimated effect sizes from the basic and fully adjusted models had a Pearson’s correlation of 0.73 at p < 2.2 x 10^-16^. Pearson correlations between cognitive outcomes and covariates ranged between -0.49 (digit symbol and age) and 0.39 (verbal fluency and vocabulary) and can be observed in **Figure S2 and Table S6**.

### Proteomic associations with *APOE* ε4 status

Using a linear mixed-effects models adjusted for age, sex and relatedness with *APOE* ε4 allele count (zero, one or two copies) modelled as a factor, we identified 11 proteomic associations (n unique proteins = 7) with carrying a single copy of the *APOE* ε4 allele (absolute β_range_ = 0.03 – 0.43, p < 3.1 x10-4) and six associations with carrying two copies (absolute β_range_ = 0.1 – 1, p < 3.1 x 10^-4^, **Figure S3, Table S7**). After further adjustment for the relevant covariates (as in the cognitive models), all Bonferroni-significant associations remained significant (absolute β_range_ = 0.03 – 1, p < 3.1 x 10^-4^, **Figure 2, Table S8**). The estimated effect sizes from the basic and fully adjusted models had a Pearson’s correlation of 0.997 at p < 2.2 x 10^-16^. Nine of these associations were with apolipoproteins. For example, carrying one or two copies of the ε4 allele was positively associated with a mass spectrometry signal that mapped to apolipoprotein B-100 (β = 0.2 and 0.3, respectively). Expectedly, carrying one or two copies of the ε4 allele was negatively associated with a mass spectrometry signal that uniquely mapped to apolipoprotein E (β = -0.4 and -1, respectively). A non-apolipoprotein, afamin, was negatively associated with carrying one copy or two copies of *APOE* ε4 (β = -0.08 and -0.2, respectively).

**Figure 2:**
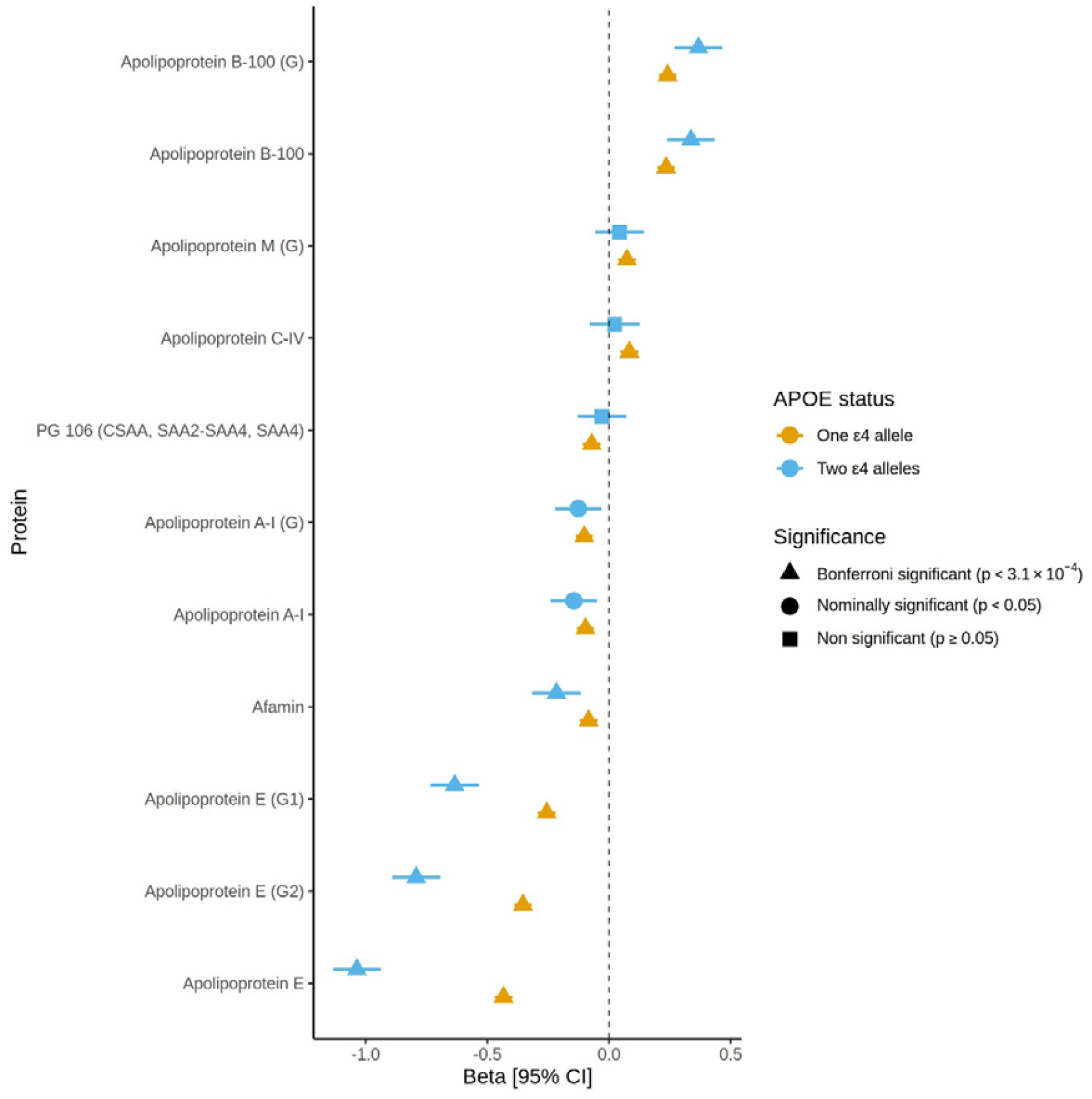
Protein/protein group associations with *APOE* ε4 status. Standardised effect sizes (Beta) for proteins/protein groups with *APOE* ε4 status in Generation Scotland for the fully adjusted models. Error bars represent 95% confidence intervals [95% CI].

**Figure 2:**
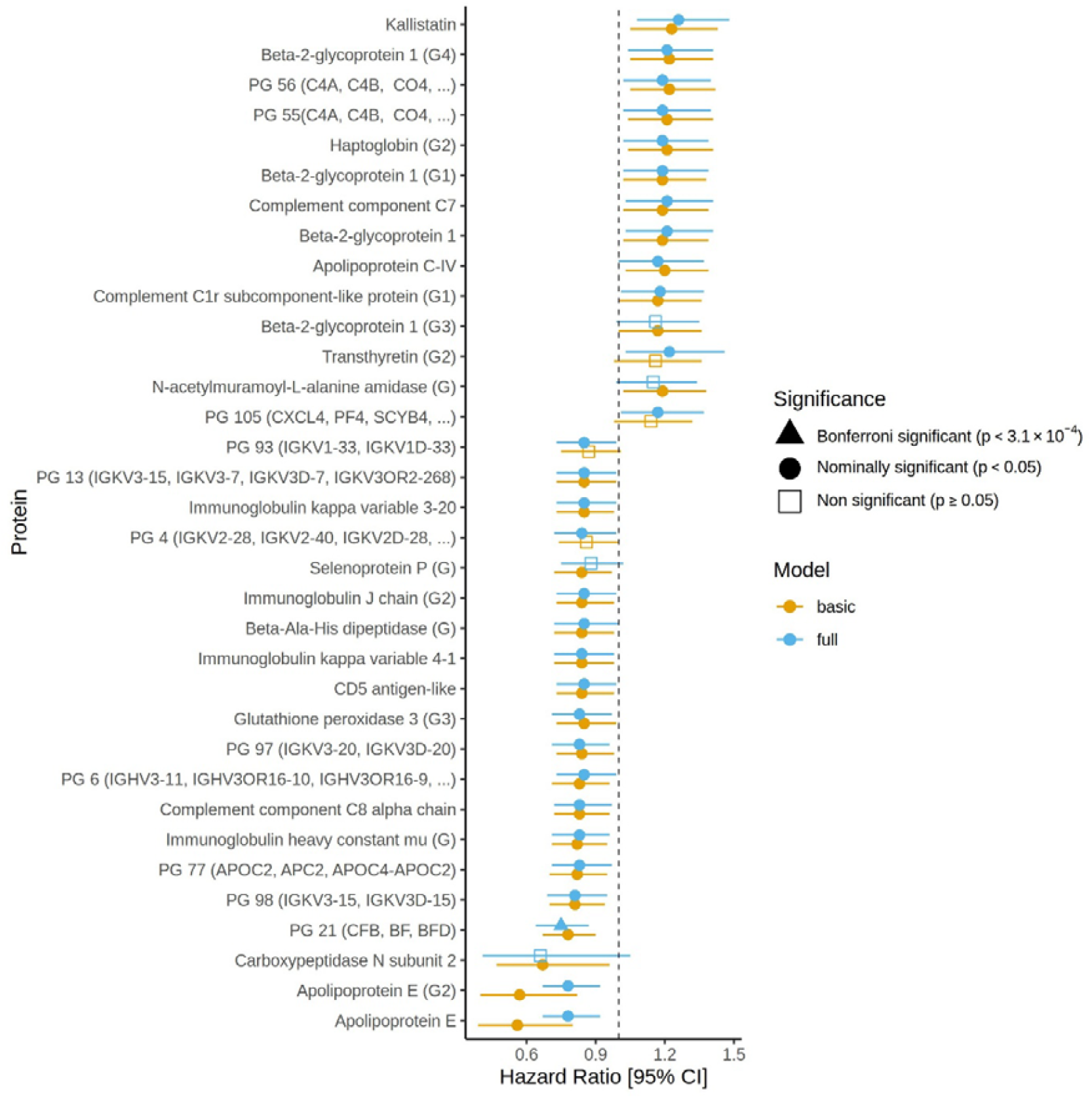
Protein/protein group associations with incident dementia in basic and full models. Hazard ratios for proteins/protein groups with incident dementia in Generation Scotland for the basic and fully adjusted models. Error bars represent 95% confidence intervals [95% CI].

### Proteomic associations with incident dementia

The relationship between incident dementia and the protein/protein group levels was explored in a subset of GS aged ≥ 65 years at diagnosis/censor using mixed-effect Cox proportional hazard models adjusted for age, sex and relatedness (n_cases_ = 216 and n_controls_ = 6,911). Thirty associations with incident dementia were identified at p < 0.05 (absolute log hazard ratio range = 0.16– 0.58, **Table S9**). No statistically significant associations were observed at a Bonferroni corrected threshold of p < 3.1 x 10^-4^. In the fully adjusted models with the additional inclusion of *APOE* ε4 status (n_cases_ = 212 and n_controls_ = 6,765), 30 nominally significant associations were found (absolute log hazard ratio range = 0.15 – 0.29, p < 0.05, **Table S10**) with incident dementia. An overlap of 26 nominally significant associations from the basic and full models was observed (**Figure 3**). A Pearson’s correlation of 0.67 at p < 2.2 x 10^-16^ was observed between the estimated effect sizes from the basic and full models. Protein group 21 (complement B factor and complement C2) was significantly associated at a Bonferroni-adjusted threshold in the fully adjusted model (hazard ratio = 0.75, p < 3.1 x 10^-4^).

## Discussion

In this study, we explored the relationship of 439 blood-based protein/protein group levels with cognitive function (n = 14,258), *APOE* ε4 status (n = 14,745), and incident dementia (n_controls_ = 6765, with n_cases_ = 212) in the Generation Scotland cohort.

We used a novel multivariate multiple linear regression Bayesian joint sparse model to simultaneously test for associations between proteins/protein groups and five cognitive function traits. Our choice of method allowed us to account for the correlation structure between proteins/protein groups whilst simultaneously considering their relationships with multiple related outcomes. Using this approach, we identified selenoprotein P (G) to be associated with three of the four cognitive tests (not logical memory) in fully adjusted models. Selenoprotein P is a protein that facilitates the transport of selenium to organs, including the brain ^34^. A recent study (n = 54) investigated the relationship between both serum and cerebrospinal fluid selenoprotein P with progression from mild cognitive impairment (MCI) to dementia (n = 35 progressing from MCI to dementia) ^35^. They found that higher levels of selenoprotein P were predictive of dementia onset, particularly when progression to dementia occurred > 2 years after baseline ^35^. Another study indicates the relationship between risk of MCI/AD with CSF/serum selenoprotein P may be an inverted U-shape ^36^. In contrast, our analysis suggests that higher levels of selenoprotein P are associated with higher cognitive functioning levels. Differences in cross-sectional vs longitudinal designs or a non-linear relationship between selenoprotein P and cognitive function could potentially explain these conflicting results.

Four associations were observed for the cognitive PGS, including complement C4-B (G), PG 52, PG 54 and PG 56 in the fully adjusted models. PG 52, PG 54 and PG 56 were all composed of various protein IDs that mapped to either complement C4-A or complement C4-B. PG 52 and PG 56 were negatively associated with the cognitive PGS, whilst complement C4-B (G) and PG 54 were positively associated. In the basic-adjusted models, complement C4-A (G) was negatively associated with the cognitive PGS, which could suggest that PG 52 and PG 56 are capturing more signal related to complement C4-A, whilst PG 54 is capturing more signal related to complement C4-B. A previous study observed a negative correlation between genetically predicted C4-A and cognition (fluid intelligence, pairs matching and digit symbol, standardised beta = 0.008, 0.006, 0.008, respectively) ^37^. We did not observe any association between C4-A (or the protein groups that include C4-A IDs) with digit symbol or any other cognitive test in our study.

Eleven proteomic associations, mapping to seven unique proteins, were observed with *APOE* ε4 status in our fully adjusted models. The majority of these associations were with apolipoproteins. The *APOE* ε4 genotype has been shown previously to associate with an increase in apolipoprotein B levels and a decrease in apolipoprotein E levels ^38–41^, as was observed in our study. In this study, afamin was negatively associated with ε4 carrier status. Afamin is a glycoprotein mainly produced in the liver that binds to vitamin E ^42^. Blood-based levels of afamin associate with several metabolic health-related disorders, including type 2 diabetes ^43,44^, gestational diabetes ^45^, non-alcoholic fatty liver disease ^46^, hepatic lipid accumulation ^44^ and metabolic syndrome ^47^. It is worth noting that the association between *APOE* ε4 status and afamin remained after adjustment for metabolic health covariates, including BMI and self-report diabetes and hypertension. Afamin levels have been shown to negatively correlate with the percentage of intermediate density lipoproteins (IDL) subfractions and mean low-density lipoprotein (LDL) size ^48^. In the same study, afamin positively correlated with the percentage of large LDL and small-dense LDL subfractions. Apolipoprotein E is involved in the clearance of IDL, and carrying two copies of the ε4 allele is associated with higher LDL ^38^. This study further supports that a relationship exists between afamin and lipid status; however, further work is required to fully understand this relationship.

In fully adjusted dementia models, we observed a Bonferroni-significant association for Protein group 21 (PG 21, hazard ratio = 0.75, p < 3.1 x 10^-4^). Five protein IDs make up PG 21. Three of the IDs (“P00751”, “H7C5H1”, “A0A0G2JH38”) are for the complement factor B protein, and the remaining two IDs (“E7ETN3”, “B4E1Z4”) relate to complement C2. Both proteins play a role in immune response via different activation pathways of the complement system ^49^. The relationship between the complement system and dementia has been explored previously ^50–53^. For example, complement C2 levels were found to be elevated in symptomatic genetic frontotemporal dementia participants (n = 104) compared with controls (n = 327) ^51^. In astrocyte-derived exosomes found in plasma, the levels of complement proteins significantly differed between individuals who transitioned from mild cognitive impairment (MCI) to dementia within three years (n = 20) compared to those with MCI that remained stable over the same time period ^50^.

A strength of this study is the modelling approach we applied to test associations between protein/protein groups and cognitive function. Firstly, by using a multivariate approach, we took into consideration the relationship between cognitive outcomes, allowing us to more accurately estimate effect sizes. Secondly, by modelling the proteins/protein groups conditionally, we account for the correlation structure between the predictors, reducing the risk of detecting false positive results. The statistical approaches for testing associations between protein/protein groups with *APOE* ε4 status and incident dementia (linear mixed effects and Cox mixed effects models, respectively) differed from the first method. It is important to note that these approaches were marginal, which could be regarded as a limitation, as they did not model the proteins conditionally, which can lead to false positive results. We aimed to mitigate this by using a Bonferroni-adjusted threshold for statistical significance, whereby we divided alpha (0.05) by the number of PCs (161) required to explain 80% of the variance in the proteomic data. This study also benefits from a large sample size and a wide age range (17-99 years) to allow for examination of biomarkers of cognitive health across the adult life-course (with the exception of the dementia analysis). As previously mentioned, most large-scale studies focus on affinity-based approaches for biomarker identification, which measure a pre-defined subset of proteins. Therefore, another strength of this study is that the proteins/protein groups were measured using an untargeted mass spectrometry approach, allowing the examination of an understudied subset of the proteome in relation to brain health. However, it is worth noting that whilst the proteins/protein groups to be measured were not pre-defined, only 439 mass spectrometry signals were reliably measured and correspond to highly abundant proteins in the blood. Our study was performed with individuals of European ancestry; therefore, future replication studies should also be performed with individuals of non-European ancestry to determine how well these biomarkers translate across populations. Finally, the current study design could not identify whether these proteins/protein groups are causally associated with cognition and dementia or a secondary change. Future work could include using Mendelian randomisation to explore if these proteins/protein groups are causal.

This work described an exploratory, discovery-stage analysis designed to identify circulating proteins associated with cognitive function, genetic susceptibility to cognitive health, *APOE* ε4 status, and future dementia risk. In this study, we have highlighted potential biological pathways and candidate biomarkers that would require replication and mechanistic follow-up to fully establish their reliability and relevance as correlates of cognitive function, genetic risk and dementia.

## Data Availability

All Generation Scotland participants signed a broad consent form. According to the terms of consent for Generation Scotland participants, access to data must be reviewed by the GS access committee. Applications should be sent to. All code for this study is available on GitHub: https://github.com/marioni-group/Mass_spectrometry_proteomics_and_cognitive_health.

