## Supplementary Figures for "Proteomic biomarkers of cognitive function, *APOE* ε4 status, and dementia in Generation Scotland"

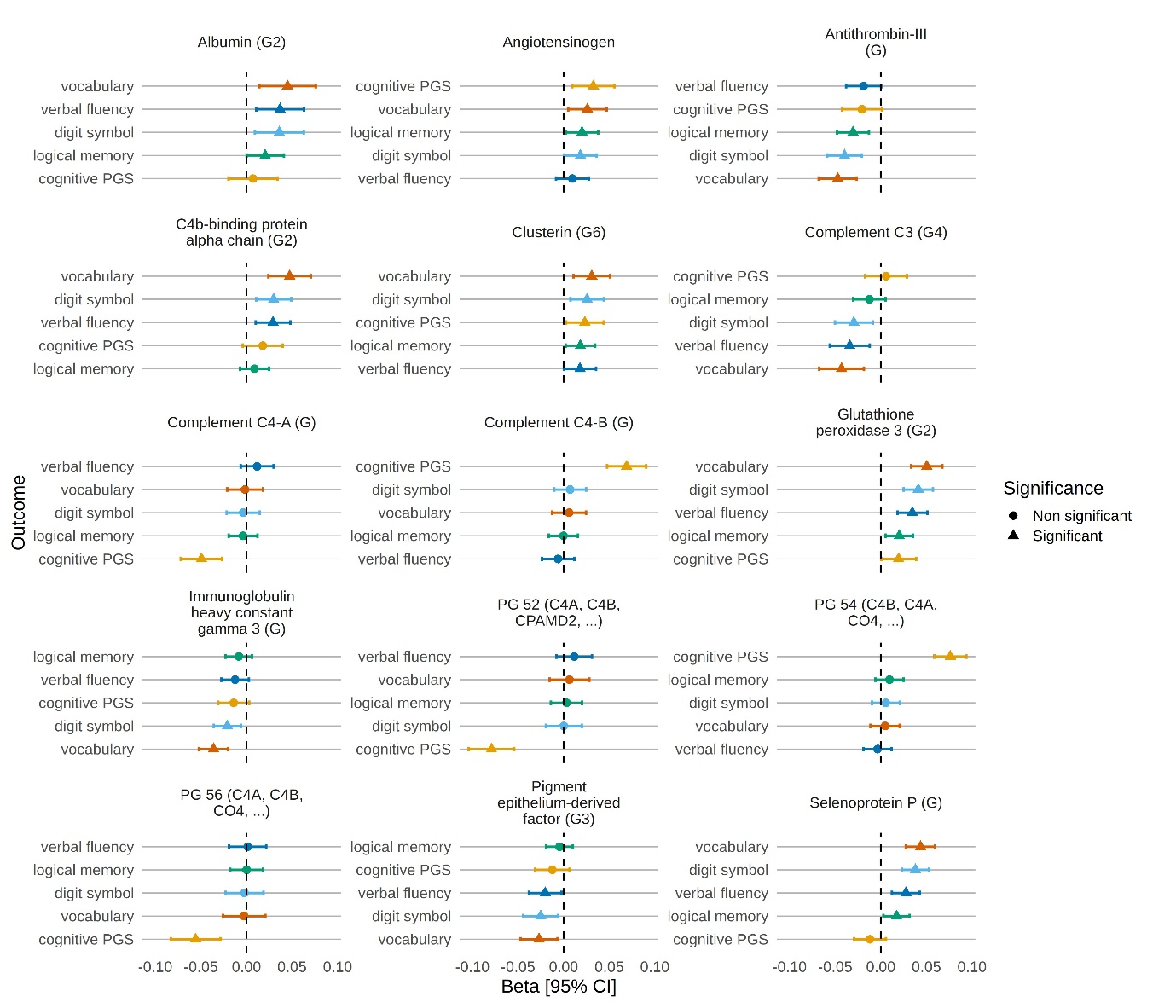

**Figure S1: Associations between cognitive outcomes and proteins/protein groups in MAJA models with basic adjustments.** Standardised effect sizes are shown for 15 proteins/protein groups with digit symbol (light blue), logical memory (green), vocabulary (orange), verbal fluency (dark blue), and cognitive PGS (yellow) in Generation Scotland. Protein/protein groups are included in the plot if they had a significant association (PIP ≥ 0.95 and the 95% credible interval did not include zero) with one or more outcomes. Error bars represent the 95% credible interval [95% CI].

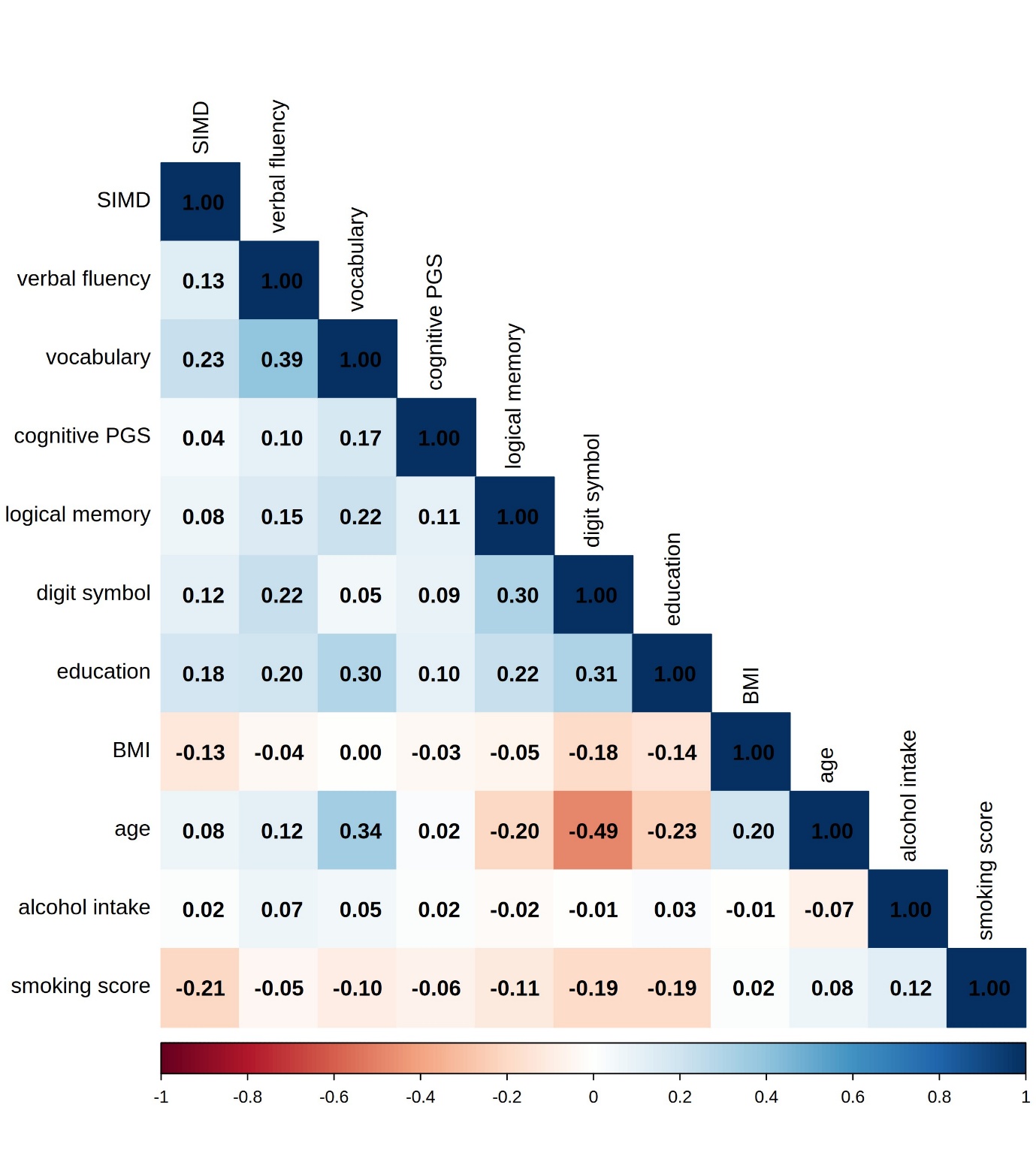

**Figure S2: Cognitive outcome and covariate correlations.** The heatmap shows the Pearson correlations between the five cognitive outcomes and all covariates used in the fully adjusted models with complete data.

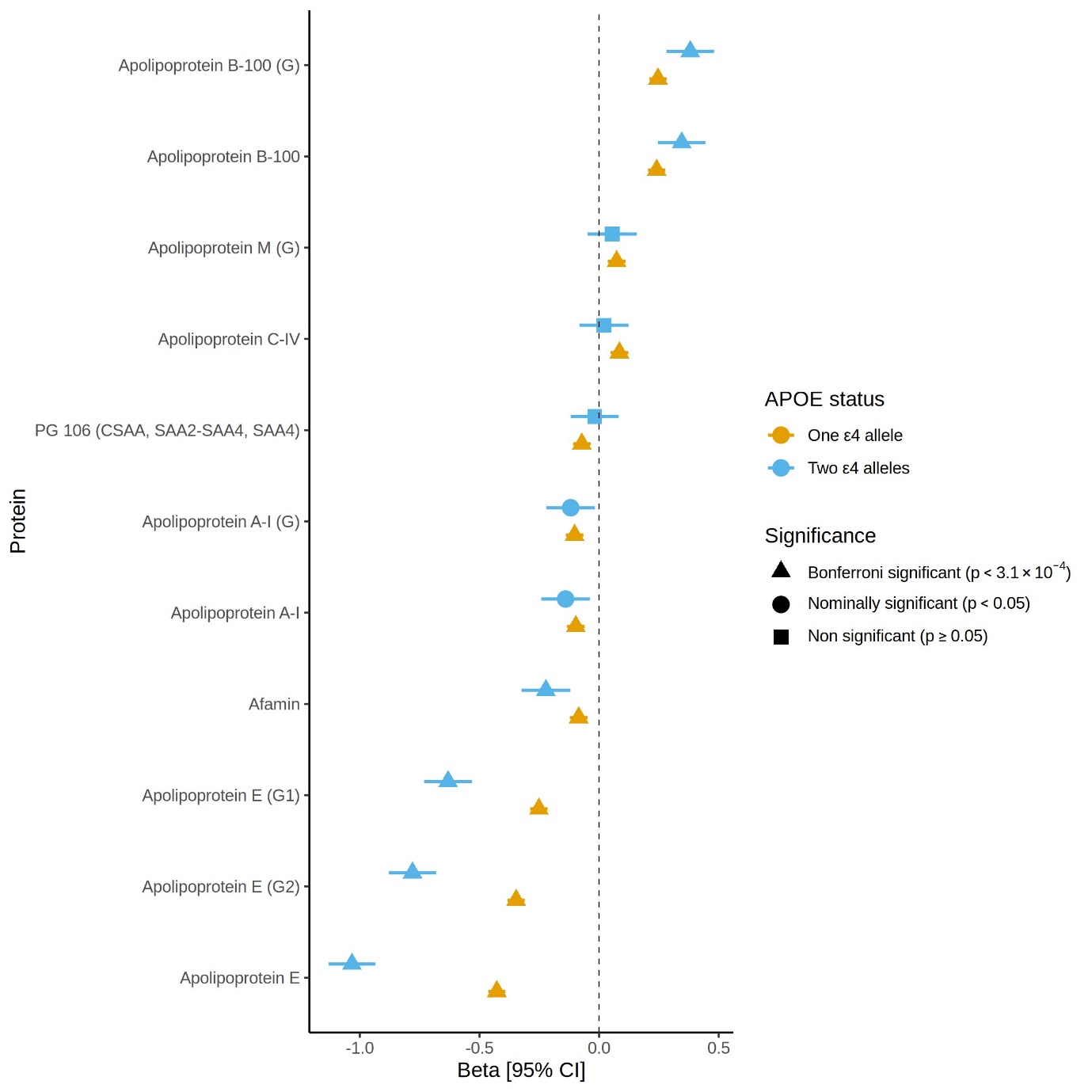

**Figure S3: Protein/protein group associations with *APOE* ε4 status**. Standardised effect sizes (betas) for proteins/protein groups with *APOE* ε4 status in Generation Scotland for the basic adjusted models. Error bars represent 95% confidence intervals [95% CI].
